# A *Trypanosoma cruzi* Trans-Sialidase Peptide Demonstrates High Serological Prevalence Among Infected Populations Across Endemic Regions of Latin America

**DOI:** 10.1101/2025.01.22.25320967

**Authors:** Hannah M. Kortbawi, Ryan J. Marczak, Jayant V. Rajan, Nash L. Bulaong, John E. Pak, Wesley Wu, Grace Wang, Anthea Mitchell, Aditi Saxena, Aditi Maheshwari, Charles J. Fleischmann, Emily A. Kelly, Evan Teal, Rebecca L. Townsend, Susan L. Stramer, Emi E. Okamoto, Jacqueline E. Sherbuk, Eva H. Clark, Robert H. Gilman, Rony Colanzi, Efstathios D. Gennatas, Caryn Bern, Joseph L. DeRisi, Jeffrey D. Whitman

## Abstract

Infection by *Trypanosoma cruzi*, the agent of Chagas disease, can irreparably damage the cardiac and gastrointestinal systems during decades of parasite persistence and related inflammation in these tissues. Diagnosis of chronic disease requires confirmation by multiple serological assays due to the imperfect performance of existing clinical tests. Current serology tests utilize antigens discovered over three decades ago with small specimen sets predominantly from South America, and lower test performance has been observed in patients who acquired *T. cruzi* infection in Central America and Mexico. Here, we attempt to address this gap by evaluating antibody responses against the entire *T. cruzi* proteome with phage display immunoprecipitation sequencing comprised of 228,127 47-amino acid peptides. We utilized diverse specimen sets from Mexico, Central America and South America, as well as different stages of cardiac disease severity, from 185 cases and 143 controls. We identified over 1,300 antigenic *T. cruzi* peptides derived from 961 proteins between specimen sets. A total of 67 peptides were reactive in 70% of samples across all regions, and 3 peptide epitopes were enriched in ≥90% of seropositive samples. Of these three, only one antigen, belonging to the trans-sialidase family, has not previously been described as a diagnostic target. Orthogonal validation of this peptide demonstrated increased antibody reactivity for infections originating from Central America. Overall, this study provides proteome-wide identification of seroreactive *T. cruzi* peptides across a large cohort spanning multiple endemic areas and identified a novel trans-sialidase peptide antigen (TS-2.23) with significant potential for translation into diagnostic serological assays.

**One Sentence Summary:** Phage display immunoprecipitation sequencing (PhIP-seq) designed with a *T. cruzi* whole proteome library reveals a trans-sialidase peptide antigen (TS-2.23) with antibody responses highly prevalent across endemic regions of Latin America.

## INTRODUCTION

Chagas disease is caused by infection with the protozoan parasite, *Trypanosoma cruzi*, which is transmitted by triatomine insect vectors. The disease is endemic to the Americas, with vector-borne transmission occurring in suitable ecological zones of Latin America (*1*). In the United States (US), the major disease burden occurs among Latin American immigrant populations exposed in their birth countries, although rare autochthonous infections have been documented in Texas, California, Arizona, Tennessee, Mississippi and other southern states (*2, 3*). Chronic Chagas disease is considered a lifelong infection without treatment. *T. cruzi* can infect many nucleated cell types but causes pathology in the cardiac and gastrointestinal systems. An estimated 20 to 30% of people with chronic Chagas disease develop symptoms of end organ damage after years to decades of infection. Related cardiac presentations include cardiac conduction system deficits, dilated cardiomyopathy, and sudden cardiac death (*4*). Ten percent of infected individuals may develop gastrointestinal dysmotility disorders (*5, 6*). Because this parasite is predominantly intracellular in the chronic phase and symptoms are largely non-existent or non-specific, detection of anti-*T. cruzi* antibodies in peripheral blood is the most sensitive method for diagnosis and is the only reliable means of screening asymptomatic patients.

The test performance of current Chagas disease serology assays does not have the accuracy (sensitivity or specificity) to effectively diagnose patients by one test alone (*7*). Pan American Health Organization/World Health Organization (PAHO/WHO) guidelines require confirmation by two tests with distinct antigen sources. The indications for *T. cruzi* serology span many areas of healthcare, including clinical diagnosis, blood donor screening, and solid organ or hematopoietic stem cell transplant donor and recipient testing (*8–11*). In practice, securing repeat testing for patients and identifying clinical laboratories that offer more than one serology test can be difficult and time consuming, and ultimately patients may be lost to follow-up. Given the mounting awareness of the need for Chagas disease screening and imperfect test performance, it is clear that the serology assays themselves must be improved to increase the effectiveness of screening and diagnosis efforts.

Recent studies evaluating regionally-diverse Chagas disease populations highlight differential reactivity to commercial *T. cruzi* serology assays between infected populations; with the lowest reactivity in individuals from Mexico, intermediate reactivity from Central America, and the highest reactivity from South America (*12–15*). Up to an estimated 10% loss in sensitivity between infections originating from Mexico compared to South America has been observed depending on the assay used (*12*). Other studies based in endemic areas have documented decreased performance of commercial serological assays in regions of Mexico and Central America, as well as Peru (*16–19*). *T. cruzi* is a genetically diverse parasite, currently classified into six genetic lineages or discrete typing units (DTUs: TcI – TcVI) (*20*), plus a potential seventh, bat-associated genotype (TcBat), most closely related to TcI (*21*). It is hypothesized that host immunological responses and antigenic differences between regional *T. cruzi* strains may be the basis of the differential serological responses. However, the areas with problematically low reactivity to commercial assays are largely found where TcI is predominant, but not all TcI-predominant areas show low reactivity (*22*). Genetic variation is also high within TcI (*23*), suggesting that DTU-level classification is not sufficiently granular to map host immunology to parasite genetics.

The antigens used in current commercial diagnostics originated from a surge of Chagas disease serology research over the last three decades (*24*). These studies tended to rely on screening with sera from high prevalence regions of South America, where TcII/V/VI are predominant; mainly Brazil and Argentina. Since then, more robust techniques for antigen discovery have emerged in the form of high-density peptide microarrays. Recent application of these techniques to Chagas disease have generated additional antigen targets (*25–27*). However, these studies used pooled sera for determining the initial down selection of antigenic targets for secondary peptide array libraries. Such an approach is unable to discern commonality of antigens across the entire *T. cruzi* proteome among the pooled sera. The antigen targets chosen for follow-on validation in individual specimens were therefore biased towards the highest reactivity antigens within a pool, not necessarily the highest prevalence antigens.

To address these gaps, we employed phage display immunoprecipitation sequencing (PhIP-seq) using a synthetic oligonucleotide library with high-density coverage of the *T. cruzi* proteome using 47 amino acid peptides. We performed immunoprecipitation using 185 serology-confirmed cases from geographically diverse regions of Latin America to explicitly represent the genetic diversity of *T. cruzi* antigens across DTUs, as well as varying presentations of Chagas cardiomyopathy to control for any differences by disease severity. The goal of this study was to employ a next-generation antigen discovery technique to *T. cruzi* and evaluate high-prevalence antigen targets with translational potential for serological diagnostics.

## RESULTS

### Development of *T. cruzi* proteome library

We constructed a T7 phage-display library to display the entire *T. cruzi* CL Brener proteome in 49-amino acid (aa) peptides with a 19-mer overlap in consecutive sequences (*Materials and Methods*, Fig. 1). The library includes 228,127 *T. cruzi* peptides that represent 19,607 proteins. Over 99.6% of the ordered peptides were represented in the final cloned library, with 90% of peptides represented within a 9-fold difference of read counts (Fig. S1).

**Fig. 1.**
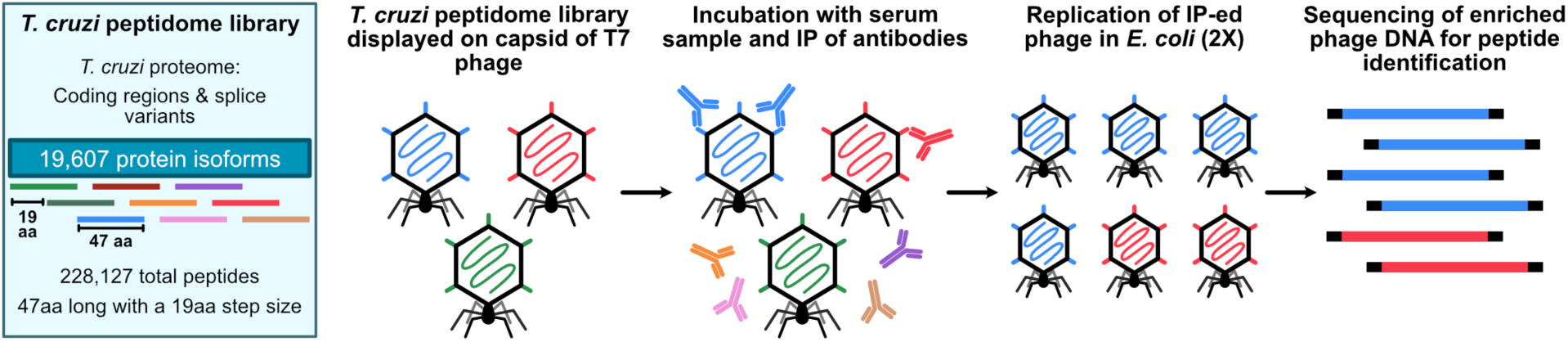
PhIP-seq library design and assay steps. Phage library displays the proteome of *T. cruzi* in 47-aa peptides with a 19-aa step size on the capsid of T7 phage. The library includes all coding regions of the proteome and splice variants. We performed the PhIP-seq assay by incubating the phage library with human plasma, followed by immunoprecipitation of antibodies in the sample and enrichment of antibody-bound phage through lysis in *E. coli*. We performed two rounds of enrichment and then sequenced the enriched phage to obtain the identity of the immunoprecipitated peptides.

### PhIP-seq identifies antibodies to *T. cruzi* peptides

We performed PhIP-seq on peripheral blood samples from three distinct specimen sets including US blood donors with routine Chagas disease serology screening (BD, *n=*90; *n*=64 seropositive, *n=*26 seronegative), a Chagas disease cardiac biomarker study (CBM, *n=*143; *n=*121 seropositive, *n=*22 seronegative), and independent healthy controls (NYBC, *n=95*). See *Materials and Methods* for a full description of these specimens. The PhIP-seq library contained human GFAP sequences, so a polyclonal anti-GFAP antibody was used as a positive control for immunoprecipitation. Positive control samples were highly enriched for GFAP peptides (Fig. S2). We excluded ten samples from the analysis due to low sequencing read counts; seven seropositive CBM samples, two seronegative CBM study samples, and one NYBC control. In total, 185 cases and 143 controls were included for further data analysis.

We used a conservative analysis approach to identify antibody reactivity to individual *T. cruzi* peptides that were enriched (z-score ≥5) in at least 5% of seropositive patients (Fig. S3). Z-scores were calculated based on the distribution of sequencing reads per 100,000 (RPK) value for a given peptide in seronegative patients, which included endemic region seronegative controls as well as independent seronegative controls from the US. With this approach, 5,638 individual peptides representing 4,001 proteins were enriched in the seropositive CBM samples, and 8,710 peptides representing 5,629 proteins were enriched in the seropositive BD samples. Between both cohorts, 12,978 antigenic peptides corresponding to 7,373 unique proteins were identified, with an overlap of 1,370 peptides across 961 proteins in both Chagas disease study sets (Fig. S4). A total of 67 and 85 peptides were reactive in at least 70% of samples among the BD samples and CBM samples, respectively. Across both cohorts, 43 of these 70% seroreactivity peptides were shared.

### Significantly enriched *T. cruzi* antigens in seropositive samples

The antigenic peptides identified across both specimen sets represented 38% of the 19,607-member proteome of *T. cruzi*. Most of these peptides demonstrated no enrichment in seronegative samples (Fig. 2a, b). The median number of enriched Chagas disease-specific peptides in each seropositive sample was higher in the BD specimens compared to the CBM specimens (Fig. 2c), but the mean number of enriched peptides per sample did not significantly differ within cohorts by patient region of origin (BD specimens) or by heart disease stage (CBM specimens) (Kruskall-Wallis, BD *H*(2) = 1.77, *p* = 0.41; CBM *H*(3) = 2.49, *p* = 0.48).

**Fig. 2.**
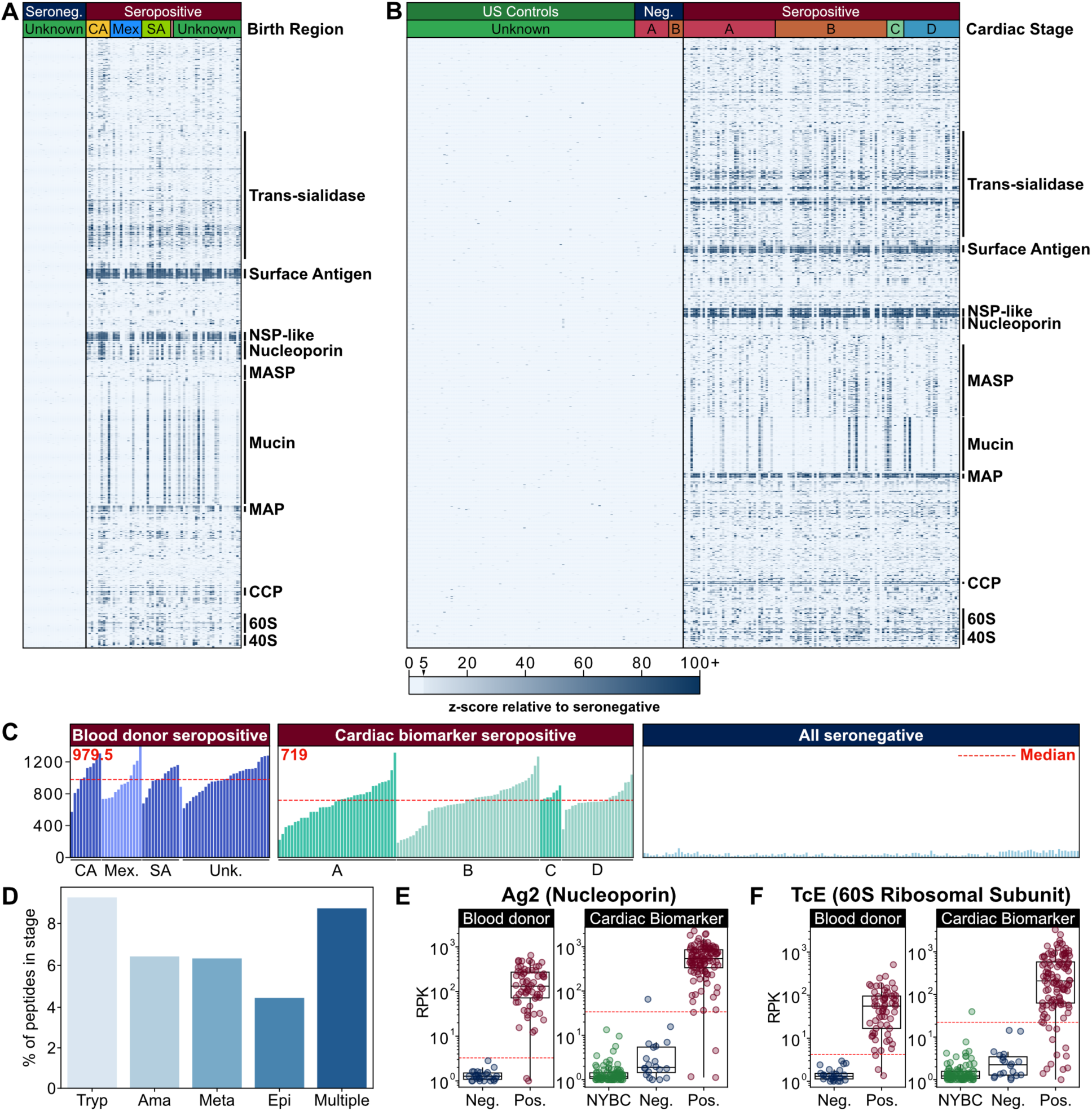
PhIP-seq captures known antigens across the *T. cruzi* life cycle stages. (**A**,**B**) Heatmap of z-score enrichment over seronegative controls in the (A) blood donor (BD) specimens (n=64 seropositive, n=26 seronegative) and the (B) cardiac biomarker (CBM) specimens (n=114, seropositive; n=18, seronegative; n=95, healthy controls) for seroreactive peptides (rows) with >15% seropositivity within each cohort. Peptides are sorted by protein name and samples are sorted by patient region of origin (n=10, Central America [yellow]; n=13, Mexico [blue]; n=12, South America [light green]; n=1, USA [pink]; n=28, unknown [green]) (A), or cardiac disease stage (n=14 stage A seronegative [magenta]; n=6 stage B seronegative [orange]; n=38 stage A seropositive [magenta]; n=46 stage B seropositive [orange]; n=7 stage C seropositive [light green]; n=23 stage D seropositive [blue]) (B). Protein groups with well-characterized antigens are indicated by labels (Surface antigen, Surface antigen 2 (CA-2); NSP-like, Nucleoporin NSP1-like C-terminal domain-containing protein; MASP, Mucin-associated surface protein; Mucin, TcMUCII; MAP, Microtubule-associated protein; CCP, Calpain-like cysteine peptidase; 60S, 40S, ribosomal subunit proteins). (**C**) Breadth of antibody reactivity, shown as the number of seroreactive peptides in each person. The dotted red line and number signify the median number of seroreactive peptides in BD and CBM specimen sets. Samples are grouped by geographic region (BD specimens) and heart disease stage (CBM specimens). (**D**) Number of peptides identified as seroreactive in this study that are part of proteins expressed in specific stages of the *T. cruzi* life cycle (Tryp = trypomastigote; Ama = amastigote; Meta = metacyclic trypomastigote; Epi = epimastigote; Multiple = protein is expressed in trypomastigote, amastigote, and/or metacyclic trypomastigote stages). Stage expression analysis shows seroreactive peptides in every host-interfacing lifecycle stage. Stage-specific expression is based on the ‘Life cycle proteome (Brazil)’ data set from TriTrypDB. Gene IDs for stage-specific proteins were mapped onto the gene IDs that corresponded to seroreactive peptides. (**E**,**F**) Selected known seroreactive antigens are captured by *T. cruzi* PhIP-seq. Neg. is seronegative specimens from the respective specimen sets; Pos. is seropositive specimens from the respective cohorts; NYBC is NYBC US controls. Antibody reactivity to two known antigens (E) Ag2, a nucleoporin protein, and (F) TCE, a 60S ribosomal subunit protein are plotted as reads per 100,000 (RPK). The dotted red line signifies the RPK that corresponds to a z-score cutoff of 5 in the seronegative population of each cohort.

The proteins from which the enriched peptides derive are predominantly expressed in the host phase lifecycle stages of *T. cruzi* (metacyclic trypomastigotes, trypomastigotes, and amastigotes) (Fig. 2d). There were relatively fewer seroreactive peptides that corresponded to proteins expressed in the epimastigote form, which only occurs in the gut of the triatomine vector. Examples of host phase-specific proteins that had high antibody reactivity included trans-sialidases and mucin-associated surface proteins.

### Identification of high-prevalence *T. cruzi* antigens across Latin America

To identify high-prevalence antigens shared across endemic regions representing different *T. cruzi* DTUs, we first analyzed BD specimens, which included individuals born in Mexico, Central America and South America. We performed two complementary approaches to identify individual antigens with sufficient seroprevalence to have utility for clinical diagnostic assays. First, we used the z-scored, peptide-level data to identify peptides enriched in ≥90% of seropositive BD samples. Second, we used mass univariate analysis to create a ranked list of top antigenic peptides by largest predictor coefficient values. The mass univariate analysis approach models peptide RPK scores based on diagnostic status in our BD specimens. All peptides identified by the z-score approach were also identified as significantly enriched by the mass univariate analysis (Fig. S5). These analyses were then performed on the CBM specimens to evaluate for any differences in high-prevalence antigens by cardiac disease status; none were identified.

These analyses yielded 23 peptides (Table S1), including 20 peptides that all contain the repetitive PFGQAAAGDKPS antigenic sequence, present in a current diagnostic antigen known as Ag 2 (*30, 31*) (Fig. 2e). An additional highly reactive peptide, which contained the antigenic sequence KAAAPKKAAAPQ, has high sequence homology to another known diagnostic antigen, TcE (*32*) (Fig. 2f). The final two high-prevalence peptides belonged to the trans-sialidase family (Fig. 3a) and shared the sequence APGETK[V/I]PSELNATIPSDHDILLEFR[D/E]LAAMALIG. To our knowledge, this peptide is novel as a diagnostic antigen candidate.

**Fig. 3.**
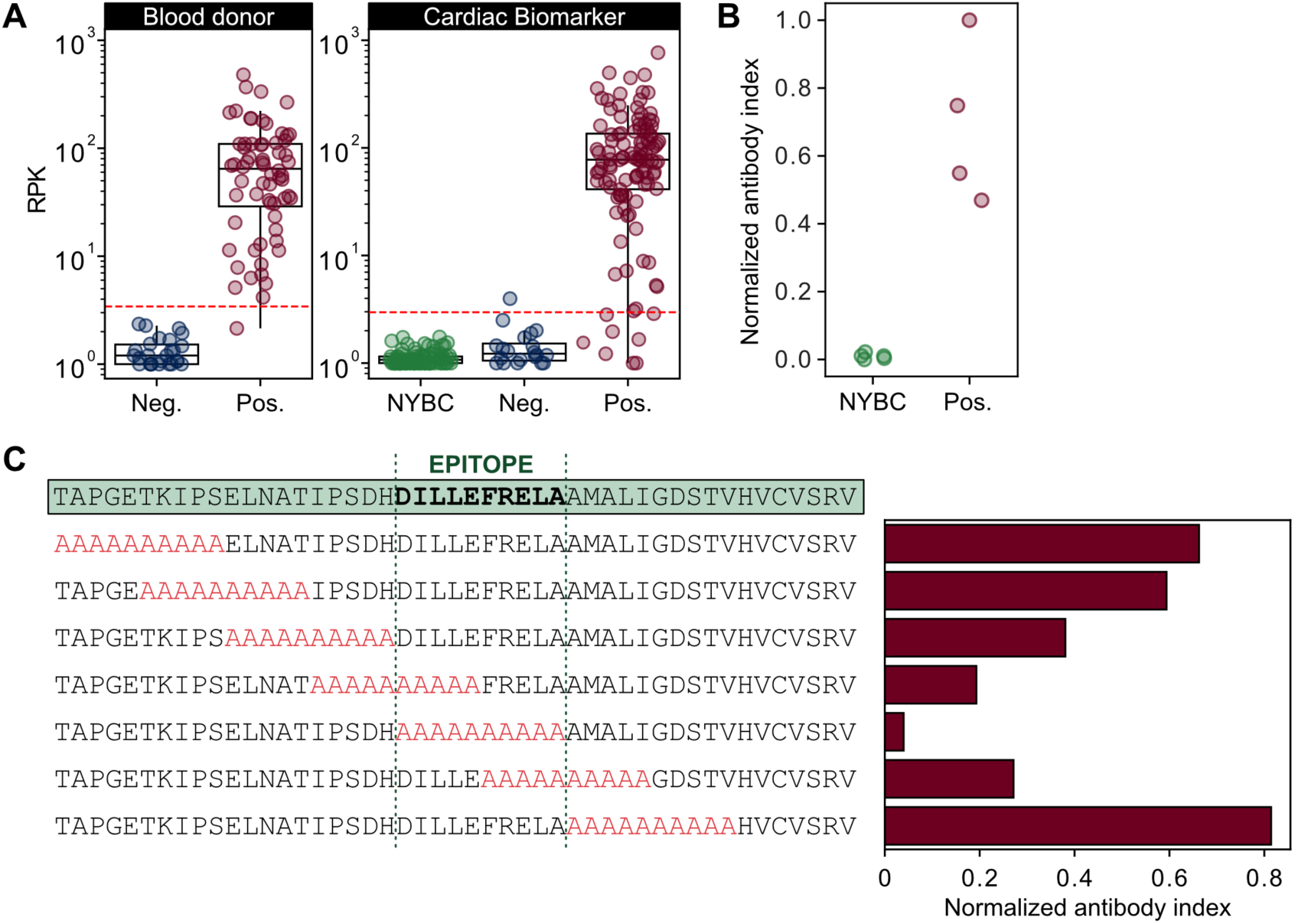
A novel trans-sialidase peptide sequence is a highly reactive serological antigen. (**A**) Anti-trans-sialidase peptide antibody reactivity is plotted as RPK. The dotted red line signifies the RPK that corresponds to a z-score cutoff of 5 in the seronegative population of each cohort. (**B**) Trans-sialidase reactivity orthogonal validation using a split-luciferase binding assay (SLBA). Reactivity was tested against four seropositive blood donor specimens and five seronegative US healthy control specimens. (**C**) Alanine-scanning mutagenesis in 10-aa windows (highlighted in red) across the entire trans-sialidase antigenic fragment demonstrates the seroreactive epitope in Chagas disease. Values are normalized antibody indices and represent the averages of five seropositive blood donor specimens.

### Epitope mapping and validation of the high-prevalence trans-sialidase antigen

To orthogonally validate antibody reactivity to the high-prevalence trans-sialidase peptide, we performed a split-luciferase binding assay (SLBA). Briefly, we generated the trans-sialidase peptide with a C-terminal HiBiT tag and immunoprecipitated it with plasma from 4 seropositive BD patient samples with the highest PhIP-seq RPK values and five independent healthy US controls. Incubation of the immunoprecipitated peptides with LgBiT produces luminescence as a quantitative measurement of antibody binding. Four Chagas disease seropositive samples with high PhIP-seq enrichment were reactive to the trans-sialidase peptide, while seronegative control samples were not (Fig. 3b).

A sequential alanine-scan was performed to further map the reactive epitope of the trans-sialidase peptide. Using samples from 5 seropositive BD patients, we determined that the critical region for immunoreactivity was a ten aa sequence that spans positions 648 to 658 of the full-length trans-sialidase protein (DILLEFRELA) (Fig. 3c). To be permissive of the epitope we chose a final antigenic sequence of IPSDHDILLEFRELA, corresponding to the two alanine blocks with the lowest reactivity, henceforth be referred to as TS-2.23. Basic local alignment search tool (BLAST) analysis of TS-2.23 in NCBI database identified 37 proteins from all *T. cruzi* entries with >93% (15-aa) sequence identity, all of which were from trans-sialidase or putative trans-sialidase genes.

### Evaluation of trans-sialidase antigen TS-2.23 antibody reactivity

To evaluate the potential of TS-2.23 as a diagnostic serology antigen, we compared its PhIP-seq performance to that of current diagnostic antigens. To address the fact that current diagnostic antigen sequences vary at certain amino acids from available sequencing data (*33*) and the specific antigen sequences used in commercial diagnostics are not publicly available for all assay, we agnostically derived the aa sequence motifs of eight diagnostic antigens used in US Food and Drug Administration (FDA)-cleared serology tests (*24*) using Multiple EM for Motif Elicitation (MEME) (see *Materials and Methods*) (Fig. S6) (*34–36*). These motifs were then queried against the entire *T. cruzi* PhIP-seq proteome using FIMO to identify all peptides with a significantly similar sequence to each antigen (*37*). The maximum z-score for each BD sample across all PhIP-seq peptides with a sequence match to a given antigen motif was plotted (Fig. 4a). The maximum z-score across all peptides with a sequence match to TS-2.23 was also shown to compare the novel antigen reactivity to those in used in current diagnostics. Any sample with a z-score of at least 5 for a peptide that contained an antigen motif was considered enriched for antibody reactivity prevalence calculations (Fig. 4b). TS-2.23 had high prevalence, demonstrated in 100% (64/64) of the seropositive BD samples and 95% (108/114) of CBM seropositive samples. By comparison, only Ag 2 and TcE had similar antibody reactivity and prevalence across seropositive specimens. In contrast, Ag 1, Ag13 and Ag36 had similar prevalence but lower reactivity, and KMP-11 was rarely enriched.

**Fig. 4.**
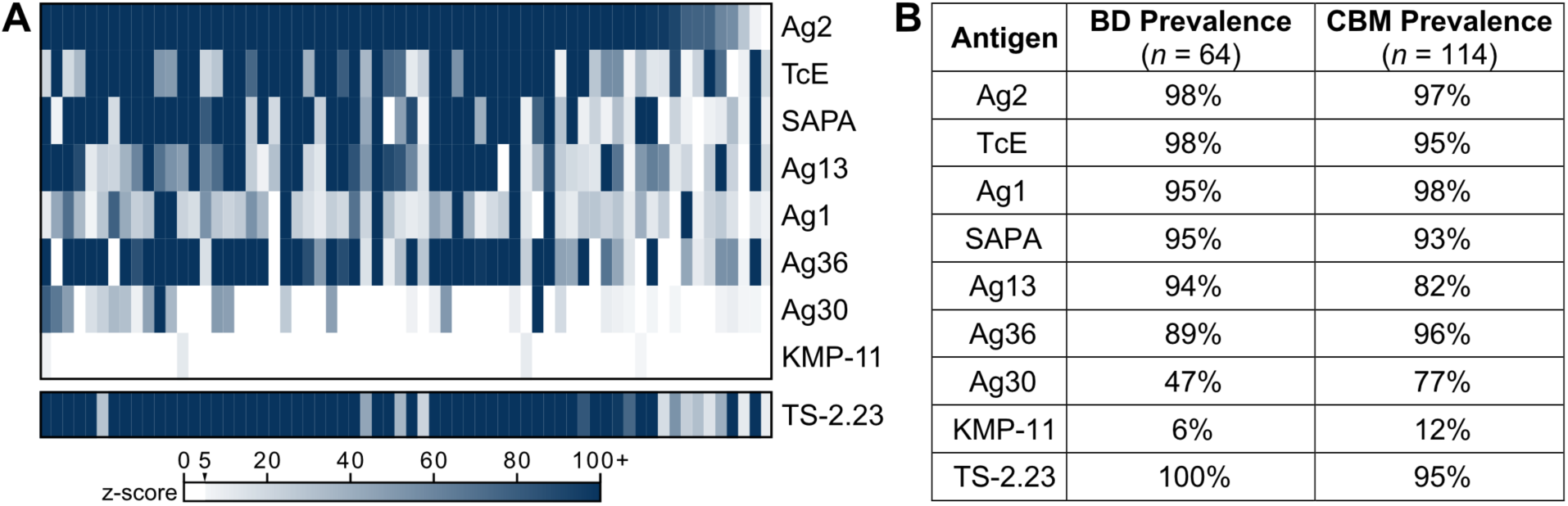
PhIP-seq antibody reactivity of current diagnostic antigens and TS-2.23 within individual Chagas disease seropositive specimens. Recombinant antigens in current FDA-cleared serology tests include Ag 1, Ag 2, Ag 13, Ag 30, Ag 36 (33, 34), shed acute phase antigen (SAPA) (35), KMP-11 (36), TcD and TcE (30, 37). Note, TcD contains the same antigenic epitope as Ag 13. (**A**) Heatmap of z-score enrichment over seronegative controls in the seropositive blood donor (BD) specimens (n=64). Each antigen motif was derived using Multiple EM for Motif Elicitation (MEME) and then scored against the entire *T. cruzi* PhIP-seq proteome. The maximum z-score across all peptides with significant sequence matches to a given antigen motif was plotted for each sample and each antigen. (**B**) Percent of samples enriched ( z-score ≥5) for each antigen in BD and cardiac biomarker (CBM) specimen sets.

To further validate and directly characterize the antibody reactivity of TS-2.23, we performed a quantitative IgG biolayer interferometry (BLI) immunoassay on 336 BD specimens with previous Chagas disease serology testing (n=250, seropositive; n=86, seronegative) (*Materials and Methods*, Fig. 5a). Seropositive samples were chosen to include all specimens with region of origin data (Mexico, n=92; Central America, n=86; South America, n=72) (Fig. 5b). The results of the pairwise comparisons between regions demonstrated seroreactivity was lower in individuals from Mexico compared to Central America (*p*<0.0001) and South America (*p*= 0.00021). Specimens from individuals from Central America and South America did not show differences in reactivity (*p*=1.000). Seronegative specimens did not demonstrate any overt non-specific reactivity to the TS-2.23 antigen.

**Fig. 5.**
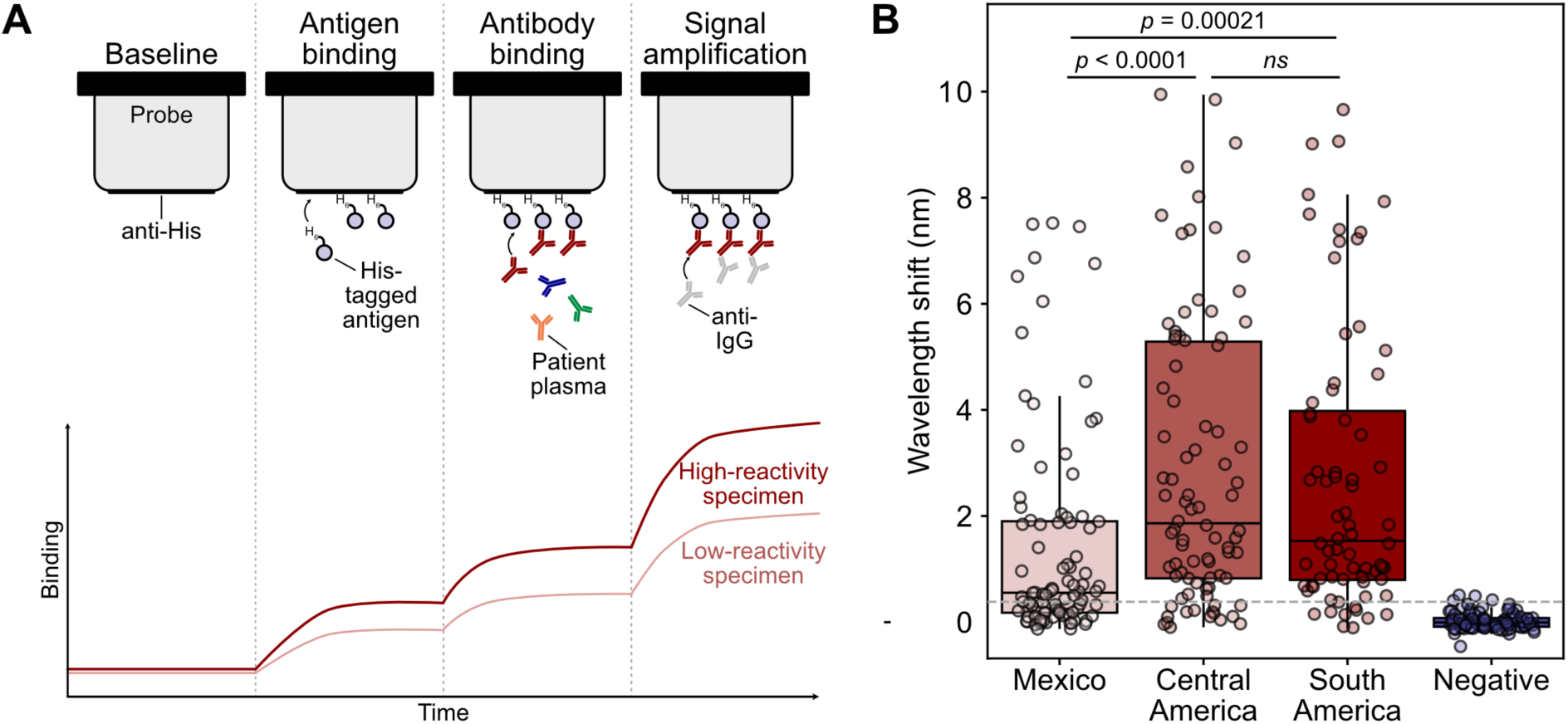
Biolayer interferometry validates trans-sialidase antigen TS-2.23 seroreactivity in a large cohort. (**A**) Schematic of biolayer interferometry (BLI) approach. BLI uses a fiberoptic probe to measure the wavelength of light reflected from the surface of a biosensor, which changes due to light interference when an analyte binds. First, an anti-His tag probe is incubated with His-tagged antigen. Then, the probe is incubated in diluted serum or plasma, and antibodies bind the immobilized antigen on the probe. To quantify IgG-specific reactivity, anti-IgG antibodies are added and bind the immobilized patient antibodies. (**B**) Seropositive blood donor specimens (n = 250) demonstrate a range of reactivity to the trans-sialidase antigen by quantitative BLI immunoassay, while seronegative blood donor specimens (n=86) do not (Wilcoxon rank-sum test). Reactivity, as denoted by wavelength shift, was higher in Central American (n=86) and South American (n=72) specimens than in Mexican specimens (n=92). Dashed line corresponds to the 25th percentile across all seropositive specimens.

To evaluate if specimens with low reactivity to TS-2.23 are weakly reactive overall, we compared TS-2.23 reactivity with previous results from an FDA-cleared Chagas disease serology ELISA (Chagatest Recombinante v.3.0, Wiener Labs [Wv3]). This assay contains a multi-epitope recombinant antigen comprised of Ag 1, Ag 2, Ag 13, Ag 30, Ag 36, and SAPA (*30, 31*). Analysis of the 25^th^ percentile of TS-2.23 BLI reactivity yielded 63 specimens originally positive by blood donor testing. The reactivity (signal-to-cutoff ratio) of the Wv3 ELISA was in the 25^th^ percentile of previous testing (*12*) for 70% (44/63) of the low reactive TS-2.23 BLI specimens. This comparison to previous Wv3 ELISA test results suggests that most low reactivity TS-2.23 specimens are weakly reactive specimens overall. Breakdown of these specimens by region of origin included 60% (38/63) from Mexico, 24% (15/63) from Central America, and 16% (10/63) from South America.

## DISCUSSION

Chagas disease is a neglected tropical disease endemic to the Americas, affecting over 6 million people worldwide (*1*). Numerous diagnostic challenges for chronic Chagas disease exist, including low clinical awareness, non-specific or asymptomatic presentation, and imperfect test performance (*38, 39*). In addition to better testing for populations with epidemiological risk factors for exposure to triatomine vectors, effective testing is needed for at-risk people presenting for prenatal screening, blood donation and organ transplant evaluation (*8–11*). The major disparities for Chagas disease serology testing are insufficient sensitivity and specificity of any one diagnostic test to be used alone and differential serological test performance by region of origin of *T. cruzi* infection.

To address this, we carried out our *T. cruzi* PhIP-seq study in two well-characterized Chagas disease specimen sets, which represent the largest number of samples evaluated for *T. cruzi* antigen discovery, to our knowledge. One set (BD) includes specimens from individuals born in Mexico, Central America and South America, collected via blood donation in the US (*12*). An additional set (CBM) includes Bolivians with varying stages of Chagas cardiomyopathy, collected via a clinical study of cardiac biomarkers (*40*). Analysis of these two specimen sets by PhIP-seq identified more than 1,300 reactive *T. cruzi* peptides highly specific to individuals with Chagas disease. Down selection on common antigens amongst Chagas disease seropositive individuals filtered to only three antigenic sequences with sufficient prevalence for diagnostic utility (enrichment in ≥90% of specimens). This vast difference between total reactive peptides and high-prevalence antigens illustrates the uniqueness of the adaptive immune system response within an individual host. By contrast, the rare high-prevalence antigens warrant further research into the host-pathogen biology that drives their immunodominance.

Two of these high-prevalence antigens were identified by multiple groups from early phage display studies and are already incorporated into current serological diagnostics: Ag 2 (*30, 31*), a nucleoporin protein, and TcE (*41*), a 60S ribosomal subunit protein. The third and, in fact, most prevalent peptide antigen discovered in our study, TS-2.23, has not previously been described as a serological diagnostic antigen and belongs to the trans-sialidase family. Trans-sialidase enzymes comprise a unique pathogenic and antigenic family of secreted and glycophosphatidylinositol (GPI)-anchored cell surface proteins in *T. cruzi*. Their primary functional activity is to transfer sialic acids from mammalian cells to beta-galactosidase residues on *T. cruzi’s* cell membrane to assist in cell invasion (*42*). Trans-sialidases also appear to modulate the host immune response, likely acting as decoy antigens (*43*). The evidence for this relates to the discovery of the shed acute phase antigen (SAPA), a multi-repeat antigen located at the C-terminal end of trans-sialidase enzymes (*44*). Since the catalytic end is in the N-terminal region, it is hypothesized that C-terminal antigens evolved to protect the trans-sialidase enzyme activity from humoral responses. BLAST analysis of the trans-sialidase antigen identified by our study showed 37 *T. cruzi* proteins with >93% sequence identity, all of which were trans-sialidase or putative trans-sialidase genes. All sequences were located at C-terminal end, matching with the immunodominant nature of known antigens from this family.

Our findings also demonstrate that the TS-2.23 antigen identified in this study has increased antibody reactivity in specimens from individuals born in Central America, compared to previous analyses of the same specimens by current clinical diagnostics (*12*). Beyond the potential implications for endemic populations in Central America, identification of TS-2.23 is important for screening and diagnosis in the US considering that a large proportion of the Latin American immigrant population is predominantly from El Salvador and Guatemala. Further studies evaluating Central American Chagas disease populations with a combination of conventional antigens and TS-2.23 will be needed to identify real-world increases in diagnostic test performance. While this is a promising advancement, unfortunately, TS-2.23 does not have improved seroreactivity in individuals who acquired *T. cruzi* infection in Mexico. Further study is needed to evaluate whether this is due to lower anti-*T. cruzi* IgG levels in individuals exposed in Mexico or the absence of regional *T. cruzi* antigens of Mexican origin. Post-translational protein glycosylation may be a source of unique antigenicity outside of the primary peptide sequence (*45*).

Our study is limited by retrospective selection bias on samples tested by current diagnostic assays. Prospective testing in at-risk populations will be important to further validate the TS-2.23 antigen. We do not report on the sensitivity or specificity of the TS-2.23 antigen in comparison to previous testing because these assays contain between 6 to 9 different antigens in a multiepitope format. The evaluation of test performance by TS-2.23 should be done in combination with these recombinant multiepitope assays to generate test performance characteristics, which are planned for follow-on studies. Ultimately, to create the ideal serologic test for diagnosis of chronic Chagas disease, we would combine the fewest recombinant targets for a multiepitope antigen that approaches 100% sensitivity and specificity to eliminate the need for confirmatory testing for all initial *T. cruzi* seropositive results. Such a test would greatly facilitate Chagas disease screening, diagnosis, and treatment.

In summary, our study has discovered a novel *T. cruzi* trans-sialidase peptide antigen (TS-2.23) that is serologically reactive and prevalent across endemic countries of Latin America and varying degrees of cardiac disease severity. Future studies will evaluate the real-world test performance in traditional immunoassay formats common in clinical laboratories.

## MATERIALS AND METHODS

### Study Design

Samples used in this study included blood donor plasma collected within the US and serum from clinical research collected in Bolivia. The blood donor plasma samples were provided in collaboration with the American Red Cross (ARC) with sample selection criteria described previously (*12*). All blood donor specimens (BD) were confirmed by blood donor testing assays and algorithms (*9*). A subset of specimens was used for antigen discovery experiments (n=90; n=64, seropositive; n=26, seronegative). Region of origin data was available for 35 specimens (n=13, Mexico; n=10, Central America; n=12, South America). The serum samples from clinical research studies in Bolivia were collected as part of a cardiac biomarker (CBM) study (*40*).

Specimens were collected from a large public hospital in Santa Cruz, tested and confirmed for *T. cruzi* serostatus and further stratified for cardiac status by clinical assessment, electrocardiogram and echocardiography studies. In total, 143 serum samples were included for this study, including 22 seronegative (n=15, without significant cardiac abnormalities [Stage A]; n=7, with cardiac abnormalities [Stage B]) and 121 seropositive (n=40, without significant cardiac abnormalities [Stage A]; n= 81, with cardiac abnormalities[Stages B-D]). Cardiac staging is defined as: A, normal electrocardiogram (ECG), normal echocardiography (echo); B, abnormal ECG, normal echo; C, 40-55% ejection fraction (EF) by echo, normal left ventricular end diastolic diameter (LVEDD); D, EF<40% or LVEDD >57mm. Specimens were randomized to 96-well plates prior to testing and frozen at -20°C. A separate set of *T. cruzi* seronegative plasma specimens (n=95) from the New York Blood Center (NYBC) was used as an independent negative control for antigen discovery experiments with the CBM specimen set.

Institutional review board for research use of de-identified human biospecimens was approved by the University of California San Francisco. The BD study protocol was approved by institutional review board at the American Red Cross. The CBM study protocol was approved by the Institutional Review Boards of Universidad Catolica Boliviana (Santa Cruz, Bolivia) and included consent for future use of deidentified specimens (*40*). NYBC specimens consisted of de-identified plasma obtained from adults who donated blood to the New York Blood Center.

### Construction of *T. cruzi* phage library

Reference protein sequences for the *T. cruzi* strain CL Brener assembly GCF_000209065.1 (*46*) were obtained from the National Center of Biotechnology Information (NCBI) site. All sequences in the peptidome were processed using a previously described bioinformatic pipeline (*47*). Briefly, all full-length protein sequences were decomposed into a series of overlapping peptides. Each peptide was 47 amino acids in length with consecutive peptides overlapping by 19 amino acids. The full set of peptides was collapsed using the command line tool cd-hit (*48, 49*) at 90% sequence similarity, resulting in a final set of 228,127 peptides spanning the *T. cruzi* peptidome (19,607 proteins). Peptides tiling over the length of the glial fibrillary acidic protein (GFAP) were added to the library as a positive control for immunoprecipitation. Peptide sequences were converted to their coding DNA sequences with common 5’ (GTAGCTGGTGTTGTAGCTGCC) and 3’ (GGTGACTACAAGGATGATGATGATAAA) linker sequences appended to each peptide encoding sequence. The 3’ linker sequence encoded a FLAG tag). The final library, consisting of 228,162 peptides that correspond to 19,608 proteins, was ordered from Agilent Technologies.

### Cloning and packaging into T7 phage

The oligo pool was received in a single tube, lyophilized, and was resuspended to 0.2 nM. The pool was amplified using Phusion polymerase (New England Biolabs [NEB]) and linker-specific primers (TAGTTAAGCGGAATTCAGTAGCTGGTGTTGTAGCTGCC, ATCCTGAGCTAAGCTTTTTATCATCATCATCCTTGTAGTCACC). The amplified library was purified using Ampure XP magnetic beads (Beckman Coulter) and confirmed to have a single-size product by gel electrophoresis. One µg of the cleaned library was then digested using EcoRI-HF and HindIII-HF restriction enzymes (NEB) and purified again using Ampure XP beads. Digestion of the library product was confirmed by visualizing a 20bp size shift using the Bioanalyzer High Sensitivity DNA Analysis kit (Agilent). The digested library was cloned into T7 Select vector arms (Novagen 70550-3) as previously described (*47*). Four packaging reactions were performed and then pooled. The final phage library was grown up in BLT5403 *E. coli* (Novagen 70550-3).

### Immunoprecipitation of antibody-bound phage

PhIP-seq was performed using the *T. cruzi* peptide phage display library with plasma or serum samples using our previously-published PhIP-seq protocol (https://www.protocols.io/view/derisi-lab-phage-immunoprecipitation-sequencing-ph-4r3l229qxl1y/v1). Patient plasma was diluted 1:1 in storage buffer (0.04% NaN3, 40% Glycerol, 40 mM HEPES (pH 7.3), 1 x PBS (-Ca and – Mg)) to preserve antibody integrity. One uL of diluted plasma was incubated with 500 µL of the input phage display library for the first round of immunoprecipitation. Positive control immunoprecipitations were performed using 1 µL of 1:10 diluted anti-GFAP antibody (Dako, Z0334) (Figure S2). Ten µL of Dynabeads Protein A/G slurry (ThermoFisher Scientific) were used per sample. After one round of immunoprecipitation, phage were amplified in *E. coli* and enriched in a second round of immunoprecipitation. The final lysate was spun and stored at 4 °C for NGS library prep. Immunoprecipitated phage lysate was heated to 70 °C for 15 min to expose DNA. DNA was then prepared for next-generation sequencing in two subsequent PCR amplifications. The final prepared libraries were sequenced using an Illumina sequencer to a read depth of approximately 1 million reads per sample.

### PhIP-seq data analysis

Sequencing reads from fastq files were aligned to the reference *T. cruzi* peptide library and individual peptide counts were normalized to reads per 100,000 (RPK) by dividing by the sum of counts and multiplying by 100,000 to account for varying read depth. All subsequent analyses were performed using Python (version 3.12.2) unless otherwise noted.

To identify Chagas disease-specific enriched peptides and avoid false positives, a conservative analysis pipeline was used as follows. Peptide-level enrichment across known seronegative samples was calculated and used to generate z-scores ((x-mean seronegative)/standard deviation seronegative) for the Chagas disease seropositive, seronegative, and NYBC control samples. The z-score for any seronegative sample was calculated by leaving out that sample from the mean of seronegative samples for each peptide. A moving threshold analysis was implemented to determine the z-score threshold and the number of Chagas disease patients that must share enrichment to a given peptide to completely differentiate seropositive and seronegative patients (Figure S3). Based on this analysis, z-score cutoff of 5 and shared enrichment across at least five percent of Chagas disease samples (n≥3 BD specimens; n≥ 5 CBM specimens) and one or fewer seronegative samples was set for hit calling.

Additional validation of the z-score approach was executed using a mass univariate analysis using generalized linear models applied to each peptide. Peptide fragments with uniform values across all samples were removed due to lack of variability. RPK values were scaled by subtracting the mean and dividing by the standard deviation calculated within each peptide.

Scaled RPK values for each peptide were regressed on Chagas disease diagnostic status (*y_i_ = β_0i_ + β_1i_ × x*) where *y_i_* is the scaled RPK value, *β_0i_* is the intercept of the *i*-th peptide fragment, *β_1i_* is the predictor coefficient, and *x* is the diagnostic status in BD samples or cardiac disease stage in CBM samples. The resulting coefficient quantified the strength and direction of the association between diagnostic status (or disease stage) and the scaled RPK values for each peptide, where positive coefficient values represent, on average, a higher RPK for that peptide in seropositive specimens. Analyses were performed using R (version 4.3.1).

Antigenic prevalence of a *T. cruzi* peptide was calculated as the number of seropositive samples enriched for a specific peptide divided by the number of seropositive samples in the respective specimen set (BD and CBM). High-prevalence antigens were designated as enrichment in ≥90% of seropositive specimens and no seronegative specimens.

### Split Luciferase Binding Assay (SLBA)

A high-prevalence antigen by PhIP-seq that was not already included in commercial diagnostics was selected for orthogonal validation by SLBA. A detailed SLBA protocol can be found online at https://www.protocols.io/view/split-luciferase-binding-assay-slba-protocol-4r3l27b9pg1y/v1. Briefly, the high-prevalence peptide antigen was inserted into a split luciferase construct containing a T7 promoter and a terminal HiBiT tag and synthesized as DNA oligomers (Twist Biosciences). The oligos were amplified using 5′-AAGCAGAGCTCGTTTAGTGAACCGTCAGA-3′ and 5′-GGCCGGCCGTTTAAACGCTGATCTT-3′ primer pair and purified using the DNA Clean and Concentrator-5 kit (Zymo). Purified PCR products were transcribed and translated *in vitro* (IVTT) using wheat germ extract (Promega L4140) and the Nano-Glo HiBiT Lytic Detection System (Promega, N3040) was used to quantify translated protein using relative luciferase units (RLU) detected on a luminometer. Background luminescence was calculated using an IVTT reaction that used a construct encoding a STOP codon 5’ of the HiBiT tag. Peptides were normalized to 2 × 10^7^ RLU per well, incubated overnight with patient plasma or a positive control anti-HiBiT antibody (Promega, N7200), and immunoprecipitated with a Dynabeads Protein A/G bead slurry. The immunoprecipitation was washed four times with SLBA buffer (0.15 M NaCl, 0.02 M Tris-HCl pH 7.4, 1% w/v sodium azide, 1% w/v bovine serum albumin, and 0.15% v/v Tween 20) and remaining luminescence was measured using the Nano-Glo HiBiT Lytic Detection System in a luminometer. Antibody index was calculated as (RLU sample – RLU mock IP)/(RLU sample – RLU anti-HiBiT) for orthogonal validation of the trans-sialidase peptides. For epitope mapping by alanine-scanning mutagenesis, the antibody index was calculated as (RLU seropositive – RLU US control)/(RLU seropositive – RLU anti-HiBiT) and normalized to the antibody index of immunoprecipitation using the wild-type peptide sequence.

### MEME and FIMO Motif Analysis

To empirically re-derive a selected diagnostic antigen motif, all BD-enriched peptides were filtered to those peptides that mapped to the antigenic protein (e.g., any enriched peptide that belonged to a nucleoporin protein for Ag2). These peptide sequences were queried using *MEME* (MEME 5.5.7) with the following *meme* command options and parameters:

- protein -mod zoops -nmotifs 10 -minw 6 -maxw 15 -objfun classic -markov_order 0

The derived motifs were then manually inspected to identify the motif that clearly matched the published diagnostic antigen sequences (Figure S6) (*24*). This motif (or multiple motifs, if the antigen sequence was over 47 amino acids, as in the case of Ag1 and Ag36) was then queried against the entire *T. cruzi* PhIP-seq proteome using the following *fimo* command options and parameters:

- -thresh 1e-4 --qv-thresh

The only exceptions to this analysis were antigens Ag13, TcE, and KMP-11. Ag13 and TcE are short, highly repetitive antigens, and so were identified using the *meme* parameter -mod anr. The final antigenic motif identified for TcE was very short (6 amino acids) and thus required different *fimo* significance thresholds to identify similar sequences. A q-value threshold of 1e-2 was set for this antigen only. Finally, KMP-11 was represented by only three overlapping peptides that map to kinetoplastid membrane protein KMP-11 (XP_808865.1), so motif discovery was not possible. To look for sequence similarity across the *T. cruzi* proteome, the 92-amino acid KMP-11 protein was queried against the proteome using *blastp* (BLAST 2.12.0) and no other peptides with significant sequence similarity were identified. The three KMP-11 peptides alone were used for downstream analysis of KMP-11 antigen reactivity.

To assess the reactivity of patient samples against these antigen motifs, the maximum z-score across all peptides with a sequence match to a given antigen motif was plotted for each BD sample.

### Peptide Antigen Expression

We selected a minimal antigenic peptide sequence that consisted of the 15-aa that, when mutated via alanine scanning, produced the lowest binding signal on SLBA (Figure 3c), to test using biolayer interferometry (BLI). This peptide sequence was repeated seven times in series to create a final protein that was approximately 13 kDa. The insert sequence was synthesized by Twist Bioscience in a pET-21(+) vector, with a C-terminal 6X His tag and under control of a T7 promoter and lac repressor.

The expression plasmid was transformed into BL21(DE3) competent *E. coli* (Thermo Scientific) and plated onto Luria-Bertani (LB) agar plates containing carbenicillin. Isolates were expanded in 1L LB broth with carbenicillin grown at 37°C to an OD600 of 0.6. The culture was induced with 1mM Isopropyl β-D-1-thiogalactopyranoside and grown at 25°C shaking for another 18 hours. The cells were then centrifuged at 10,000 RPM for 30 minutes at 4°C to collect the cell pellet.

A stock lysis buffer (20 mM sodium phosphate, 20 mM imidazole, 500 mM NaCl, 0.5 mM TCEP, 5% glycerol, pH 7.4) was made with EDTA-free (Roche) per 50 mL. The pelleted cells were resuspended in 100mL of cold lysis buffer and run through a LM10 microfluidizer at 15,000 PSI for 5 cycles. The flowthrough lysate was collected after each cycle and combined. The lysate was centrifuged at 12,500 RPM for 30 minutes at 4°C. The supernatant was collected and filtered through a 0.22 µm vacuum filtration device.

Recombinant His-tagged antigen was purified from the filtered lysate using a Ni-NTA resin gravity flow column. After loading the lysate to the column, the column was washed with a wash buffer (20 mM sodium phosphate, 40 mM imidazole, 500 mM NaCl, 0.5 mM TCEP, pH 7.4). The antigen was eluted with an elution buffer (20 mM sodium phosphate, 500 mM imidazole, 500 mM NaCl, 0.5 mM TCEP, pH 7.4). Peptide yield from the purification was quantified using NanoDrop (Thermo Scientific), and the purity of the product was verified by protein gel electrophoresis. Expression of the peptide was confirmed by anti-His tag Western blot using a 6X-His tag monoclonal antibody (Invitrogen, MA1-21315).

### Biolayer Interferometry (BLI) Serological Immunoassay

A GatorPrime analyzer (Gator Bio) was used to perform BLI to evaluate the antibody reactivity to the recombinant peptide antigen. BLI uses a fiberoptic probe to measure the wavelength of light (nanometers [nm]) reflected from the surface of a biosensor, which shifts in response to analyte binding (Figure 5a). Quantitative BLI serological immunoassay can be performed by measuring nm shift to antigen-bound probe incubated in diluted serum or plasma and subsequently in anti-human immunoglobulin (IgG) for quantifying class-specific responses. BLI methodology was chosen for these analyses because it has a higher dynamic range for assessing antibody-antigen reactivity compared to traditional colorimetric enzyme-linked immunosorbent assays (ELISA) (*50*). An anti-*T. cruzi* IgG BLI method was developed using a commercial *T. cruzi* Chimeric Chagas Multi-Antigen (MACH; Jena Biosciences). This is a polypeptide chain of 87-aas with epitopes from previously known antigens: Peptide 2, TcD, TcE, and SAPA, fused with a 6His-Tag. This BLI method was optimized using high, intermediate, and low reactivity seropositive BD specimens previously determined by Chagatest Recombinante v.3.0 anti-T.cruzi ELISA (Wiener Labs), which contain the MACH antigens.

The anti-*T.cruzi* IgG BLI assay was adapted for the recombinant antigen discovered by PhIP-seq by varying the protein concentration to achieve saturation of nm shift signal of the anti-His tag fiberoptic probe (Figure S7). The final method consisted of the following BLI conditions: 1) 600 second (s) incubation of anti-His probe in 2ug/mL peptide antigen, 2) 1800s incubation in 10uL of plasma diluted 1:19 with Q-Buffer diluent (GatorBio), and 3) 2000s incubation in a solution of 10ug/mL goat anti-human IgG (Jackson Immunoresearch). Steps 1 and 2 were followed by a 360s wash in Q-Buffer. Endpoint nm shift measurements were normalized by subtracting the nm shift value after antigen loading wash (step 1) to account for any minor variation in the amount of immobilized antigen.

Anti-*T. cruzi* IgG BLI was performed on 336 BD specimens (n = 250, seropositive; n=86, seronegative) to evaluate antibody reactivity to the peptide antigen. Region of origin data was available for all seropositive specimens (Mexico, n=92; Central America, n=86; South America, n=72). Wilcoxon rank sum analysis with a correction for multiple comparisons using the Bonferroni method was completed to compare reactivity between regions.

### Statistical analysis

Associations between number of individual antibody targets and heart disease stage or region of infection were tested using Kruskal-Wallis tests. Motif analysis was performed using MEME and FIMO (*34–36*). Associations between anti-TS-2.23 BLI reactivity, serologic status, and region were tested using the Wilcoxon rank-sum test with a correction for multiple comparisons using the Bonferroni method.

## Data Availability

All data produced in the present study are available upon reasonable request to the authors. Complete data will be made available via a Dryad repository upon peer-reviewed publication.

## Acknowledgments

We thank members of the DeRisi Lab for helpful discussions during these studies. We also acknowledge the New York Blood Center for contribution of healthy control plasma. Contents herein are the sole responsibility of the authors and do not necessarily represent the official views of the NIH or other funding agencies.

## Author contributions

Conceptualization: JLD, JDW, CB, JVR

Methodology: JDW, JLD, CB, JVR, HMK, RJM, EDG, NLB, JJP, WW, RLT, SLS, EEO, JES, EHC, RHG, RC

Investigation: HMK, RJM, JDW, JVR, AM, NLB, GW, AM, AS, CJF, EAK, ET

Formal analysis: HMK, RJM, JVR

Visualization: HMK, RJM, JDW, JVR

Funding acquisition: JLD, JDW

Project administration: JDW, JLD

Supervision: JDW, JLD, CB, EDG

Writing – original draft: JDW, HMK, RJM

Writing – review & editing: All co-authors

## Data and materials availability

All raw and processed data will be available for download on Dryad. PhIP-seq analytical code will be available at https://github.com/hkortbawi.

