## Supplementary Figures for "A *Trypanosoma cruzi* Trans-Sialidase Peptide Demonstrates High Serological Prevalence Among Infected Populations Across Endemic Regions of Latin America"

**SUPPLEMENTARY MATERIALS**

**Title:** A *Trypanosoma cruzi* Trans-Sialidase Peptide Demonstrates High Serological Prevalence Among Infected Populations Across Endemic Regions of Latin America
SHORT TITLE: *Trypanosoma cruzi* Diagnostic Antigen Discovery

**Table of Contents:**

**Figure S1:** Histogram of read counts of the *T. cruzi* proteome PhIP-seq library

**Figure S2:** Glial fibrillary acidic protein (GFAP) enrichment is specific to polycolonal antibody control samples

**Figure S3:** Empirical determination of optimal threshold for calling peptides as seroreactive

**Figure S4:** Overlap in seroreactive peptides in blood donor (BD) and cardiac biomarker (CBM) specimen sets

**Figure S5:** Mass univariate analysis of PhIP-seq results

**Figure S6:** Antigen motifs

**Figure S7:** Empirical determination of antigen concentration for optimal biolayer interferometry (BLI) performance

**Table S1:** Ranked list of *T. cruzi* PhIP-seq antigenic peptides by prevalence of enrichment in BD specimens

**Authors:**
Hannah M. Kortbawi[1,2]†, Ryan J. Marczak[3,4]†, Jayant V. Rajan[1‡], Nash L. Bulaong[5], John E. Pak[5], Wesley Wu[5], Grace Wang[5], Anthea Mitchell[5], Aditi Saxena[5§], Aditi Maheshwari[3,4||], Charles J. Fleischmann[4¶], Emily A. Kelly[4], Evan Teal[4], Rebecca L. Townsend[6], Susan L. Stramer[6††], Emi E. Okamoto[7#], Jacqueline E. Sherbuk[7]**, Eva H. Clark[8,9], Robert H. Gilman[10], Rony Colanzi[11], Efstathios D. Gennatas[3, 12], Caryn Bern[3]*, Joseph L. DeRisi[1,4]*, Jeffrey D. Whitman[3*]

†, Authors contributed equally.
‡, JVR current affiliation, Current affiliation Pfizer, Inc; Collegeville, PA, USA.
§, AS current affiliation, Department of Immunology and Infectious Diseases, Harvard T. H. Chan School of Public Health; Boston, MA, USA.
||, AM current affiliation, Keck School of Medicine, University of Southern California, Los Angeles, CA, USA.
¶, CJF current affiliation, Noorda College of Osteopathic Medicine; Provo, UT, USA.
#, EEO current affiliation, Independent Consultant.
**, JES current affiliation, University of South Florida; Tampa, FL, USA.
††, SLS current affiliation, Infectious Disease Consultant, North Potomac, MD

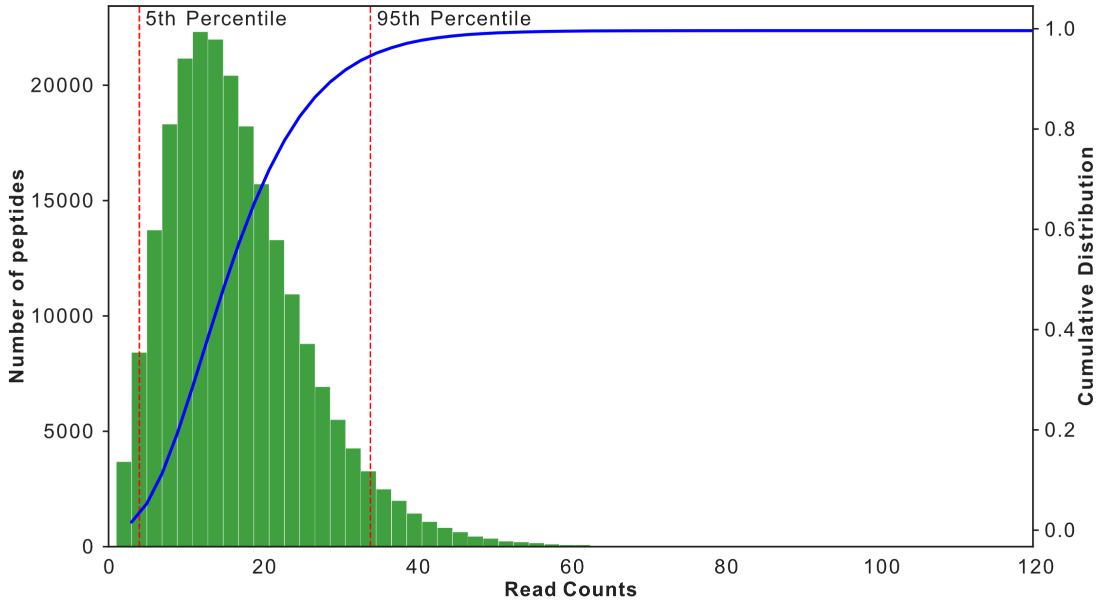


**Figure S1. Histogram of read counts of the *T. cruzi* proteome PhIP-seq library**. Read counts corresponding to the 5th and 95th percentile in the distribution (marked by red dotted lines) are within 9-fold of each other. Cumulative density plot of the distribution is shown in blue.

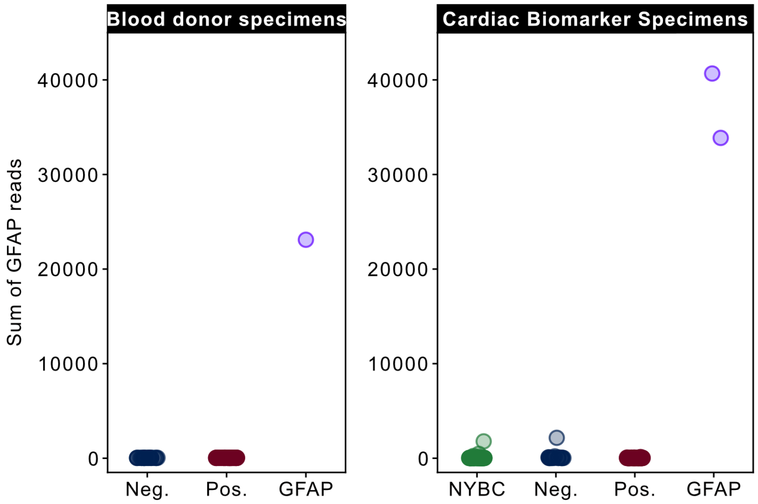


**Figure S2. Glial fibrillary acidic protein (GFAP) enrichment is specific to polycolonal antibody control samples.** GFAP enrichment is plotted as the sum of read counts for all peptides in the *T. cruzi* PhIP-seq library that are part of the GFAP protein. In both PhIP-seq runs, only sample wells with polyclonal antibody to GFAP showed increased GFAP reads.

### Blood donor specimens

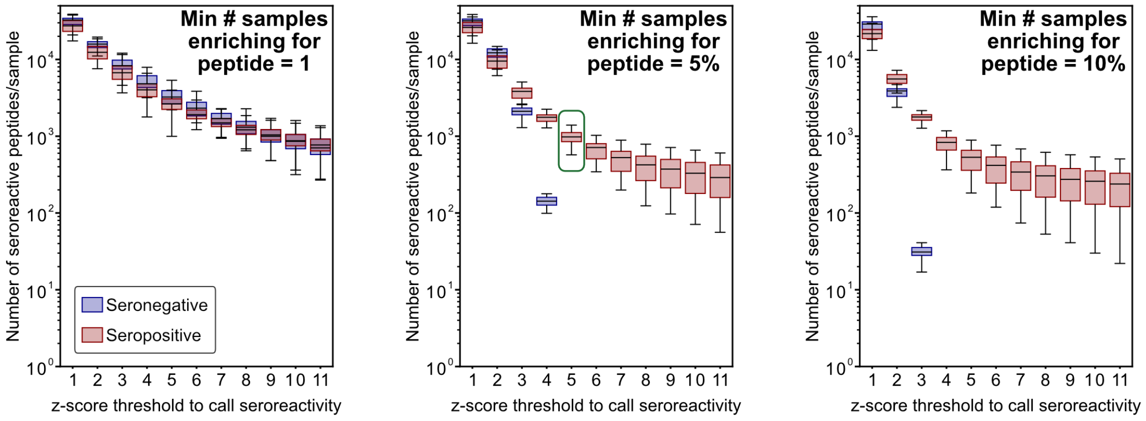


### Cardiac Biomarker cohort

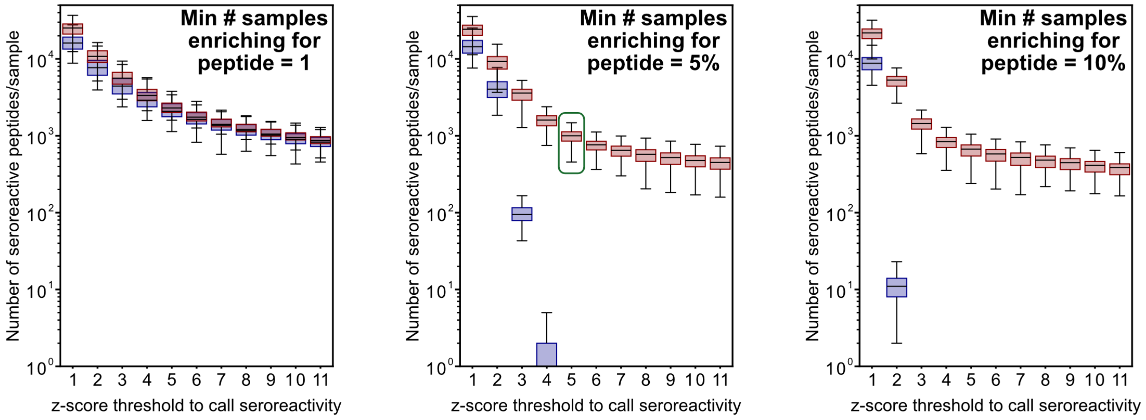


**Figure S3. Empirical determination of optimal threshold for calling peptides as seroreactive.** Box plots showing the number of seroreactive peptides per specimen at each threshold of antigen sharing (n=1, 5%, or 10% of seropositive specimens) and at z-score cutoffs of 1-11 over the mean in seronegative specimens. The final threshold for calling seroreactivity is indicated by the green boxes and was selected to minimize the number of seroreactive peptides in seronegative samples.

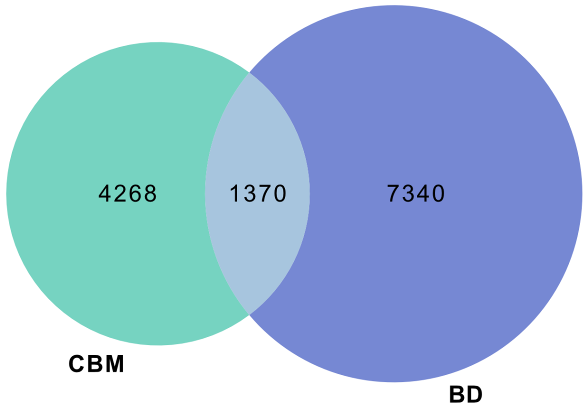


**Figure S4. Overlap in seroreactive peptides in blood donor (BD) and cardiac biomarker (CBM) specimen sets**.

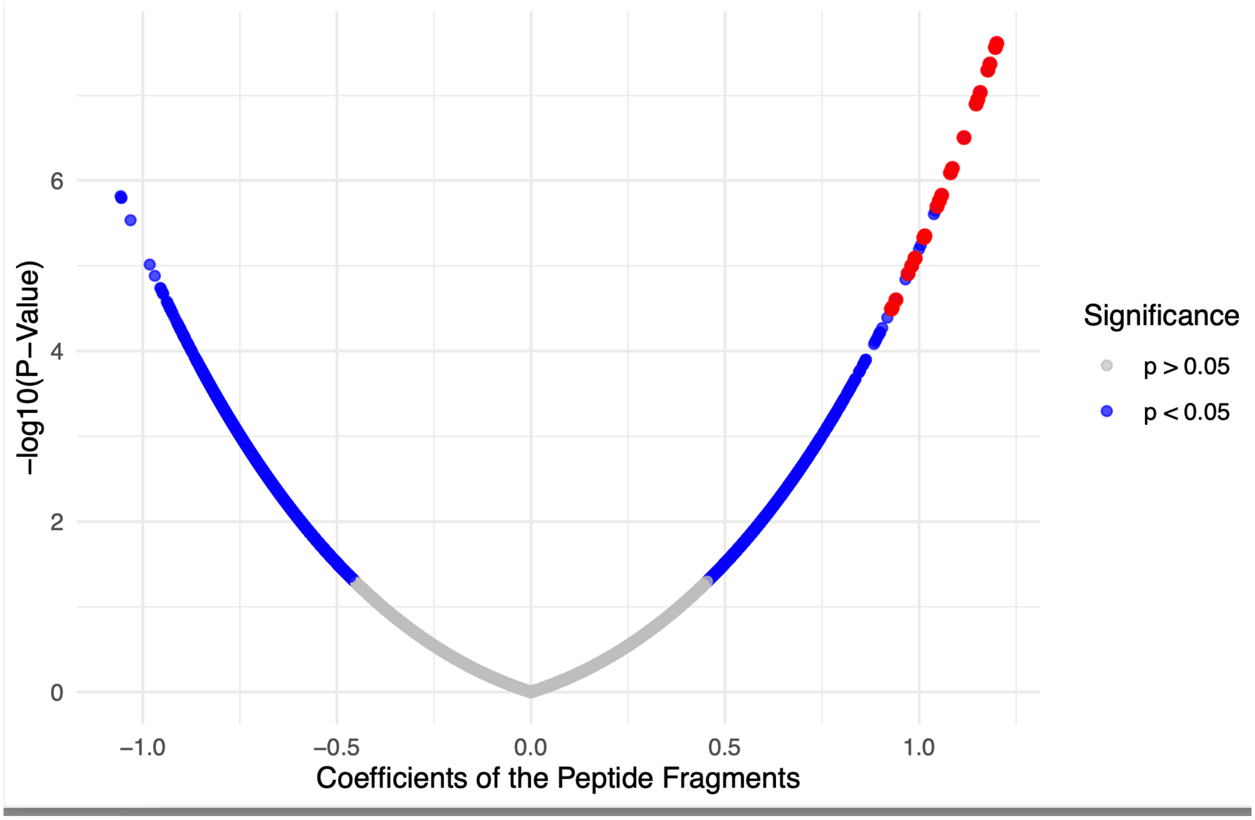


**Figure S5. Mass univariate analysis of PhIP-seq results.** This volcano plot shows the results of mass univariate analysis. The coefficient refers to the term given by the linear regression result. Coefficients>0 are associated with positive specimens. Each dot represents a single peptide, and blue dots have a *p*-value < 0.05. Red dots represent peptides that were identified as seroreactive in ≥90% of BD specimens by z-score analysis and *p* < 0.05 by mass-univariate analysis (n=23 peptides). A ranked list of peptides is available in Table S1.

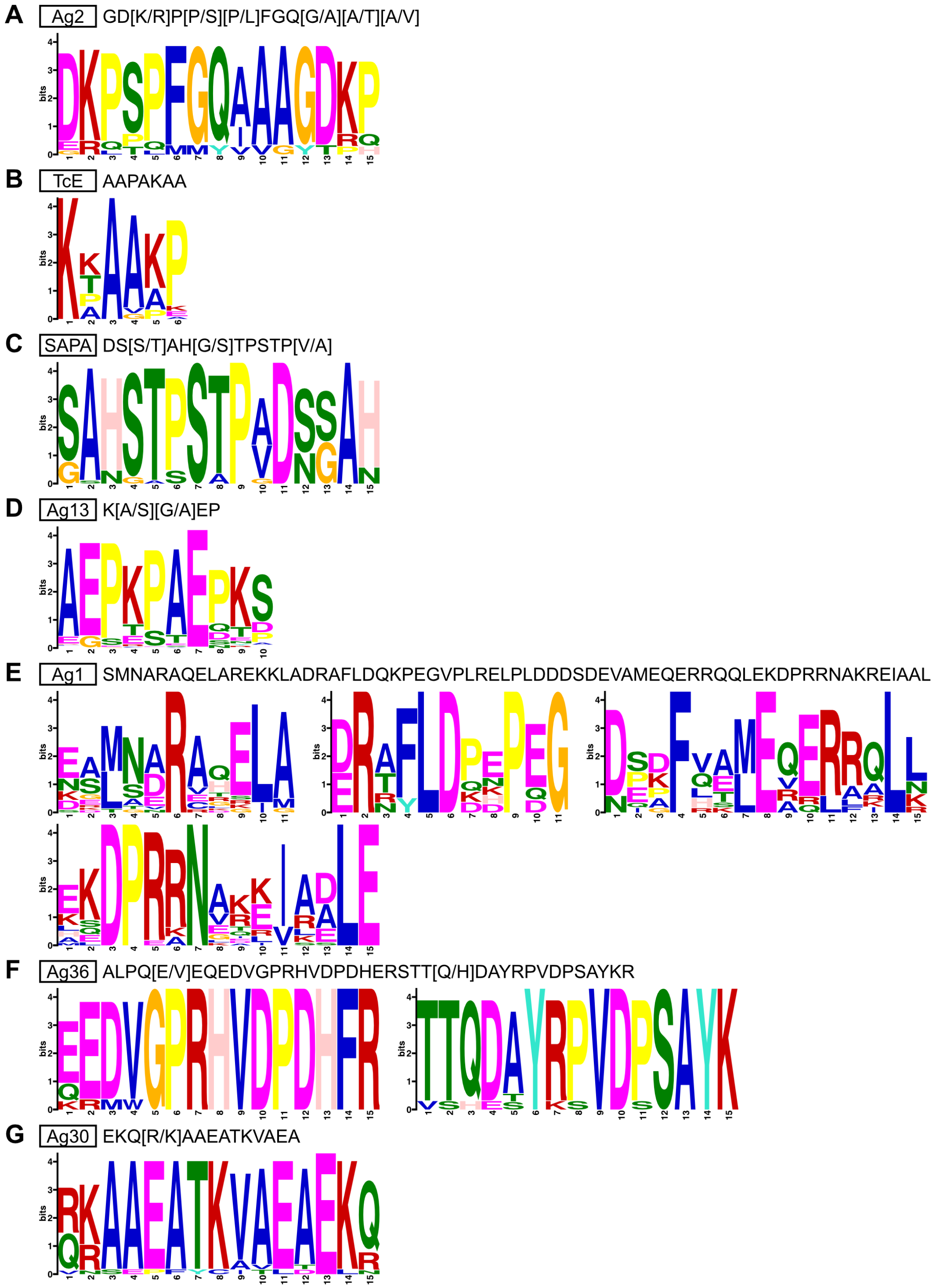


**Figure S6. Antigen motifs.** Multiple EM for Motif Elicitation (MEME) analysis was performed on seroreactive peptides that mapped to a known antigen protein (e.g., all nucleoporin peptides were analyzed to find the Ag2 motif). The output motifs are shown here, with the reference sequence (*24*) for the given antigen in plain text above the motif diagram.

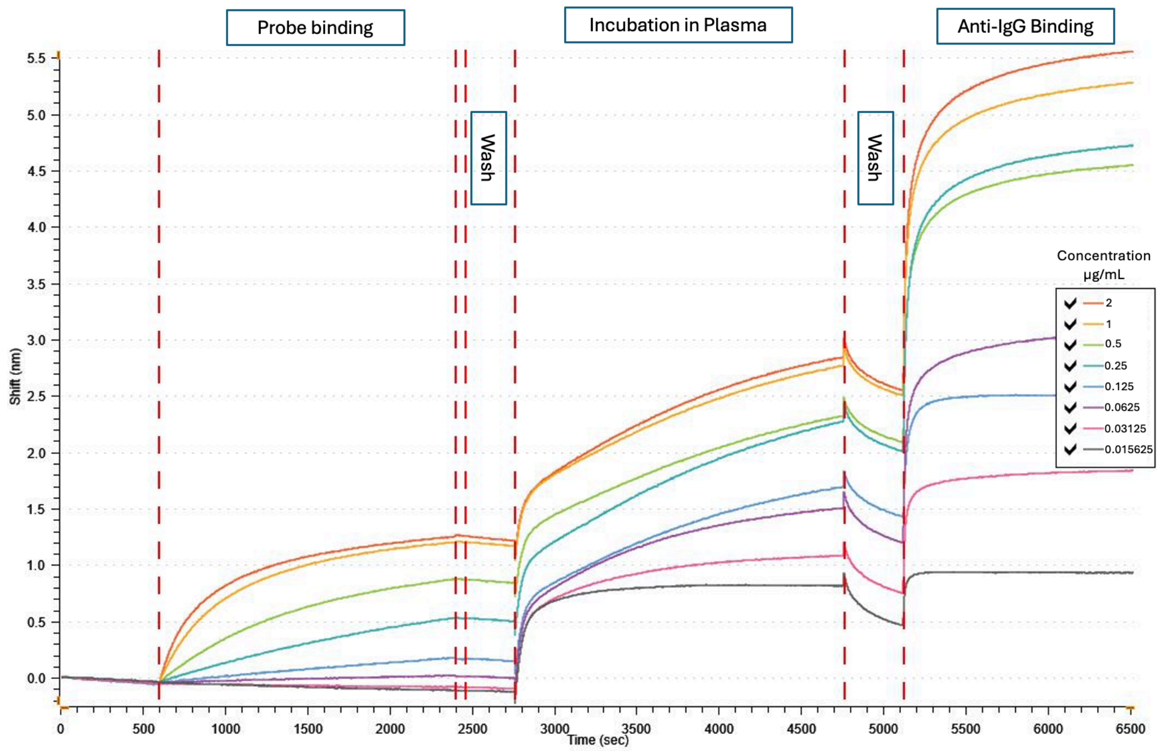


**Figure S7. Empirical determination of antigen concentration for optimal biolayer interferometry (BLI) performance.** Eight protein concentrations were tested, ranging from 0.015625 µg/mL to 2 µg/mL. 2 µg/mL was selected as the optimal concentration for further assays.
